# Imputing cognitive impairment in SPARK, a large autism cohort

**DOI:** 10.1101/2021.08.25.21262613

**Authors:** Chang Shu, LeeAnne Green Snyder, Yufeng Shen, Wendy K. Chung, on behalf of the SPARK Consortium

## Abstract

**Background:** Diverse large cohorts are necessary for dissecting subtypes of autism, and intellectual disability is one of the most robust endophenotypes for analysis. However, current cognitive assessment methods are not feasible at scale.

**Methods:** We developed five commonly used machine learning models to predict cognitive impairment (FSIQ<80 and FSIQ<70) and FSIQ scores among 521 children with autism using parent-reported online surveys in SPARK, and evaluated them in an independent set (n=1346) with a missing data rate up to 70%. We assessed accuracy, sensitivity and specificity by comparing predicted cognitive level against clinical IQ data.

**Results:** The elastic-net model has good performance (AUC=0.876, sensitivity=0.772, specificity=0.803) using 129 predictive features to impute cognitive impairment (FSIQ<80). Top ranked predictive features included parent-reported language and cognitive levels, age at autism diagnosis, and history of services. Prediction of FSIQ<70 and FSIQ scores also showed good prediction performance.

**Conclusions:** We show cognitive levels can be imputed with high accuracy for children with autism, using commonly collected parent-reported data and standardized surveys. The current model offers a method for large scale autism studies seeking estimates of cognitive ability when standardized psychometric testing is not feasible.

**Lay summary:** Children with autism who have more severe learning challenges or cognitive impairment have different needs that are important to consider in research studies. When children in our study were missing standardized cognitive testing scores, we were able to use machine learning with other information to correctly “guess” when they have cognitive impairment about 80% of the time. We can use this information in research in the future to develop more appropriate treatments for children with autism and cognitive impairment.

## Introduction

There is significant phenotypic and genetic heterogeneity in ASD that complicates the ability to identify etiologies. The need for larger sample sizes for subtyping has led to the call for large-scale studies using remote phenotyping protocols (Warnell et al., 2015, Wang et al., 2016, Samocha et al., 2014). However, current clinical assessment methods are not feasible at scale.

IQ (intelligence quotient) is a key phenotype in autism research, for stratification of the population into subtypes, prediction of individual outcomes, and correlation with genes and genetic architecture (Munson et a., 2008; Mason et al., 2020; Anderson et al., 2014; Gradzinski et al., 2013; Allegrini et al., 2019; Benyamin et al., 2014, Davies et al., 2011, Steffanson et al., 2014; Robinson et al., 2017, Ronemus et al., 2014, Rauch et al., 2012; Iossofiv et al., 2014; De Rubeis et al., 2014; Sanders et al., 2015; Grove, 2019; Satterstrom, 2020). For children with autism, formal cognitive assessment requires expert in-person examination that is costly, not equally accessible to all, and not always feasible in children with the most severe forms of autism (Corona, et al., 2021). In lieu of full assessments, proxy measures of IQ, such as receptive vocabulary and single visual reasoning tasks are used (Hamburg et al., 2019; Frazier et al., 2004; Kriseleva et al., 2017, Dawson et al., 2007), and efforts are underway to develop web-based cognitive testing applications (for example, Hansen, 2016; Scott et al., 2020). However, these testing methods are not yet fully validated for remote in-home use either in children or in individuals with autism, as would be required for large studies, nor are they appropriate for very young children or severely impaired individuals who require significant support for online tasks. Sometimes the Vineland Adaptive Behavior Scales are used as a proxy for cognitive ability (Charman et al., 2011); however, its scores show non-linear associations with IQ due to discrepancies in daily functioning, particularly for those with higher intellectual ability (Bolte and Poustka, 2002; Freeman et al., 1998; Alvares et al., 2020). Clinicians and researchers have used parent report of estimated developmental ages and intelligence, but the accuracy of this method is inconsistent (Chandler et al., 2016). Ideally, clinical IQ test data could be abstracted from medical or school records, but the resources required to do this across a national cohort is challenging (Coleman et al., 2015; Maenner et al., 2020).

As an alternative, methods for estimation or imputation of cognitive level based on IQ may be used. When IQ scores are not available, cognitive ability is often imputed by alternative measures such as educational attainment in teens or adults using educational records (Cornish et al., 2015). This approach is not possible for many autistic individuals whose educational history follows a different path such as therapeutic or ungraded settings, especially in children. When only a small proportion of participants are missing IQ scores, multiple imputation can be used to impute IQ scores (Horwood et al., 2018; Reid et al., 2018). However, this method does not work well on data with significant missingness. Alternatively, missing cognitive test scores can be imputed by linear regression models based on scores from other related concurrent measures, but the performance of these models typically has not been examined in terms of sensitivity or specificity (for example, see Emerson et al., 2016).

Linear regression also has been used to predict IQ in typical children using available demographic information, such as age, sex, parent education, and family literacy level (Schoenberg et al. 2007). Traditionally, imputation with regression models typically involves a priori subjective judgments for a small number of predictors within large datasets. Its prediction performance may be limited due to missing potentially critical variables unknown to the investigators, and the subjective selection itself can lead to overfitting and less generalizability to other datasets. When dealing with complicated and large datasets, it may have limited performance when compared to other advanced methods such as classification and regression trees (Finch et al., 2011). Studies predicting psychological traits increasingly use machine learning models. Machine learning techniques do not require a priori selection of predictors, are able to uncover latent and complicated relationships between a large number of data points, and emphasize the evaluation of prediction accuracy. Most uses of machine learning for cognitive estimation have involved prediction of future cognitive status in other clinical groups, such as older adults, (Graham et al., 2020), and although machine learning and clustering models are being applied to autism, they are largely used for trait-based subtyping and detection (Lombardo, Meng-Chuan & Baron-Cohen, 2019; Veatch et al., 2013).

SPARK (SPARK Consortium, 2018; Feliciano et al., 2019) offers the opportunity to leverage a large, heterogeneous autism cohort with phenotypic data collected online, along with clinical IQ data available for a subset of participants. It enables us to develop and assess machine learning prediction models for predicting cognitive level, using commonly collected parent-reported information. We hypothesized that cognitive impairment and IQ scores in children with ASD can be predicted based on parent-reported information such as medical and developmental history and standardized questionnaires. We developed machine learning models to impute cognitive impairment using both a binary classification model with different IQ cutoffs and a continuous model, with comparison to traditional methods such as multiple imputation. The current study seeks to fill the gap in available methods to assess cognitive impairment in autism, for use in large scale autism and autism genetics research studies.

## Methods

### 1. Sample overview

The SPARK cohort is currently the largest genetic study of autism. Participants are recruited through a network of US clinical sites and social media, and the study is conducted entirely online. As of early 2021, there were 84,161 children and dependents with a parent-reported professional ASD diagnosis (**Figure 1**). IQ scores were collected on 3,136 participants with ASD, and 2,545 participants have full scale IQ (**Figure 1**). We selected 521 participants with a complete dataset for initial model development, and another newly collected independent set (n=1,346) to further assess the model’s performance (**Figure 1**).

**Figure 1:**
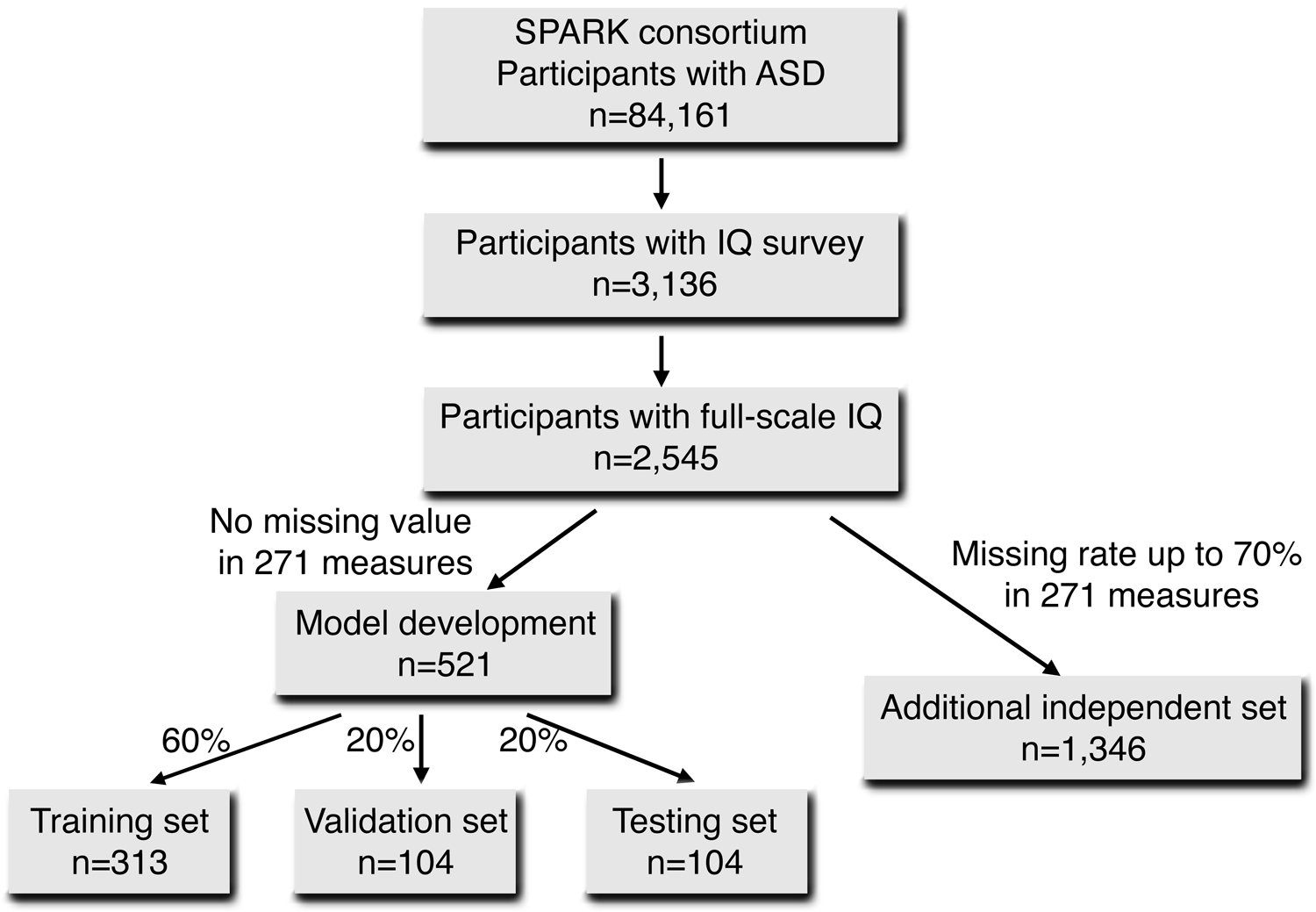
Sample selection for 521 participants with complete data for model development including training, validation and testing set. An additional independent set with 1,346 participants with missing rate<70% among 271 features was also selected

### 2. Measures

Surveys of demographics, diagnostic details, medical and developmental history, and standardized instruments including the Social Communication Questionnaire-Lifetime (SCQ-L; Rutter et al., 2003), Developmental Coordination Disorder Questionnaire (DCDQ; Wilson et al., 2007), Repetitive Behavior Scale-Revised (RBS-R; Bodfish et al, 2000), and Vineland 3rd Edition Parent-Caregiver Comprehensive rating form (Sparrow et al., 2016) were completed by parents online. Item level data and total scores were utilized for analyses. At enrollment, caregivers reported any history of ID and current functional language level (nonverbal/single words/ phrases/complex sentences).

Parents were asked questions regarding developmental milestones and regression (language and/or other skills), prior cognitive testing results, estimates of functioning, and special education services and therapies. The medical history collects data on major co-morbid medical, developmental and psychiatric conditions.

Estimated cognitive functioning level was reported by parents in several ways: overall IQ testing score if available, the mental age (age equivalent) they had been given by clinical or school providers, and general cognitive level (degree of delay) relative to chronological age. IQ test scores and mental age equivalents reported by parents could not be utilized due to missing data, which reflects parent uncertainty or lack of access to test results.

### 3. Key IQ indices collected at clinical sites

Full Scale IQ (FSIQ), Nonverbal IQ (NVIQ) and Verbal IQ (VIQ) data were collected by research staff from review of electronic and paper clinical and research records at 24 SPARK clinical sites. Staff were trained in data abstraction and required to complete and submit Case Report Forms to demonstrate inter-rater agreement with their site supervisor (psychologist), which was required to be 100% in order to proceed with data collection. Recent reports of inter-rater reliability in SPARK were high (ICC=0.98) for abstraction of IQ data using these methods (Fombonne et al., 2021). Participants age 15 months and older were prioritized, given more limited confidence in diagnosis and test results in infancy. FSIQ, NVIQ, VIQ, and developmental quotients in the case of young children, were accepted for a specific list of common, standardized, norm-referenced tests. Testing was the most recent evaluation on record. IQ assessment data were accepted if they were verified by site staff to be provided by a qualified professional (licensed M.A. or Ph.D.) in psychology or by a psychometrician supervised by a licensed psychologist, including clinical, school or research evaluations. To standardize collection of IQ data across sites, a training manual and online data entry form containing validators were provided. Any concerns regarding validity of the test results by the original test examiner of record also were entered as a flag, and such cases were excluded from analyses. All participants signed authorization within the SPARK informed consent permitting bidirectional data sharing of IQ and other information between SPARK staff and clinical sites.

IQ test data included but were not limited to the Wechsler Intelligence Scales for Children (Wechsler, 2014), Stanford-Binet Intelligence Test (Roid, 2003), Mullen Early Learning Composite (Mullen, 1995), Bayley Scales of Infant Development Mental Development Index and Cognitive Composite (Bayley, N., Aylward, G.P., 2019), and the Differential Ability Scales Global Cognitive Ability (Elliott, 2007). The indices accepted were measurements scaled to a mean standard score of 100 and standard deviation of 15. Not included were school readiness screeners, achievement tests, parent-reported developmental screeners or adaptive functioning interviews/surveys. Questionnaire measures were taken within a median of 16 months from the IQ test date among all participants.

### 4. Statistical association between IQ domain scores and between IQ scores and other covariates

IQ scores were coalesced across test types based upon their common scaling (Mean=100, Standard Deviation=15) as is common in autism research given the flexible assessment approaches required for valid comparisons (Bishop et al., 2015). Pearson correlations were calculated between FSIQ, NVIQ and VIQ scores. Pairwise scatterplots with a fitted linear model were used to assess whether the relationships are linear.

Similarly, we also assessed the relationship between IQ scores and age of IQ testing by Pearson correlation and scatterplots. Lowess smoothing was used to illustrate the relationship between IQ scores and age when IQ score was tested.

We also assessed the association between IQ scores and other variables available in SPARK, including those known to be strongly and consistently predictive of IQ across age in autism (Simonoff et al., 2019) - reported ID, estimated cognitive level (above, at or below chronological age level), language level at enrollment, history of any developmental delay and parent-report of motor.

### 5. Machine learning methods to predict cognitive impairment

FSIQ was used to define cognitive impairment since the largest number of records were available and it is highly correlated with NVIQ and VIQ. We defined cognitive impairment as FSIQ<80, in order to 1) increase the likelihood of identifying all individuals with subaverage intellectual functioning for genetic studies, 2) align better with current diagnostic criteria for ID which do not require an IQ cutoff of 70, and 3) prevent the inappropriate classification of average IQ individuals together with individuals with borderline intellectual functioning (Munson et al., 2008; Nouwens et al., 2017). The goal of the machine learning model is to distinguish participants with FSIQ <80 versus FSIQ ≥80. Because FSIQ<70 also is commonly used to define cognitive impairment, we applied the same model training, selection and testing procedures to develop an elastic net model to predict participants with FSIQ<70. Sensitivity analysis was conducted using FSIQ <70/FSIQ ≥ 70 and continuous prediction of FSIQ. There were 271 predictive measures included. Vineland scores were not included due to a high missing rate.The full list of final selected predictors is included in **Supplementary Table 1**.

a. Training, validation and testing sets The data in the initial sample (n=521) were randomly split into 60% training (n=313), 20% validation (n=104) and 20% testing data (n=104) while keeping the proportion of lower IQ consistent across training, validation and testing sets (**Table 1**). The model training was conducted in the training set, the validation set was used to select the best performing model, and the testing set was used to decide the predictive probability cutoff and model evaluation. An evaluation was also conducted to assess the model performance in the presence of missing values (up to 70%) in an additional independent set (n=1,346; **Figure 1, Table 1**).
b. Model training, selection and evaluation In the training set, we trained the following commonly used machine learning models for classification using the R package *caret* Version 6.0-88 (Kuhn, 2008): elastic-net regularized generalized linear models with no interaction terms (glmnet), support vector machines (svmRadial), random forest (rf), k-Nearest Neighbors (kNN) and gradient boosting (xgbTree). These five models are commonly used in solving classification problems (Friedman, 2001; Maglogiannis, 2007; Ogutu, 2011; Chen, 2016). The elastic-net model is a modified version of linear regression with a penalty on the number of covariates, and is easy to interpret. Support vector machines, random forest, k-nearest neighbors and gradient boosting trees can deal with high-dimensional datasets but are often hard to interpret. Ten-fold cross-validation was used in the training to minimize overfitting. Model parameters were chosen by a random grid of 50 possible combination of values and optimal parameters were chosen by the best receiver operating characteristic (ROC) among cross-validated resamples in the training set. As a sensitivity analysis for predicting continuous FSIQ, only elastic-net, support vector machines and k-Nearest Neighbors were suitable for continuous prediction and were used in the training. In the validation set, we evaluated the prediction performance for the five trained models by receiver operating characteristic (ROC) curve and area under the curve (AUC). The model with the highest AUC was selected as the final model. As a sensitivity analysis for predicting continuous FSIQ, we used the Spearman correlation between true and predicted FSIQ, root mean square error (RMSE), Cohen’s kappa, sensitivity and specificity. In the testing sets, we further evaluated the prediction performance of the final model by ROC curve, AUC, accuracy, Cohen’s kappa, sensitivity, specificity and positive predictive values (PPV) and negative predictive values (NPV). We determined the predictive probability cutoff for cognitive impairment in the initial testing set.
c. Variable importance ranking The variable importance was obtained by elastic-net model using 100 bootstraps conducted with the R package *caret*. The variable importance was scaled from −100 to 100 with absolute value reflecting the relative importance and directionality reflecting the direction of predicting low IQ.

**Table 1:** Sample characteristics

|  | Model development (n=521) |  |  |  |  |  |  |  | Additional independent set<br>(n=1,346) |  |
| --- | --- | --- | --- | --- | --- | --- | --- | --- | --- | --- |
|  | Overall |  | Training |  | Validation |  | Testing |  | Count (%) | Mean (SD<sup>†</sup>) |
|  | Count (%) | Mean (SD<sup>†</sup>) | Count (%) | Mean (SD<sup>†</sup>) | Count (%) | Mean (SD<sup>†</sup>) | Count (%) | Mean (SD<sup>†</sup>) |  |  |
| Total Sample Size | 521 |  | 313 |  | 104 |  | 104 |  | 1346 |  |
| Age At Registration in Years (mean (SD)) |  | 7.93 (3.27) |  | 7.88 (3.18) |  | 8.00 (3.32) |  | 8.01 (3.53) |  | 8.63 (6.44) |
| Sex at birth (%) |  |  |  |  |  |  |  |  |  |  |
| Female | 98 (18.8%) |  | 62 (19.8%) |  | 18 ( 17.3%) |  | 18 ( 17.3%) |  | 289 (21.5%) |  |
| Male | 423 (81.2%) |  | 251 (80.2%) |  | 86 (82.7%) |  | 86 (82.7%) |  | 1057 (78.5%) |  |
| Race (%) |  |  |  |  |  |  |  |  |  |  |
| African American | 37 ( 7.1%) |  | 27 ( 8.6%) |  | 5 ( 4.8%) |  | 5 ( 4.8%) |  | 78 ( 5.8%) |  |
| Asian American | 9 ( 1.7%) |  | 5 ( 1.6%) |  | 2 ( 1.9%) |  | 2 ( 1.9%) |  | 21 ( 1.6%) |  |
| Native American | 1 ( 0.2%) |  | 1 ( 0.3%) |  | 0 ( 0%) |  | 0 ( 0%) |  | 2 ( 0.1%) |  |
| White | 377 (72.4%) |  | 218 (69.6%) |  | 79 ( 76.0%) |  | 80 ( 76.9%) |  | 746 (55.4%) |  |
| More Than One Race | 90 (17.3%) |  | 57 (18.2%) |  | 17 ( 16.3%) |  | 16 ( 15.4%) |  | 165 (12.3%) |  |
| Other | 7 ( 1.3%) |  | 5 ( 1.6%) |  | 1 ( 1.0%) |  | 1 ( 1.0%) |  | 60 (4.4%) |  |
| Autism Diagnosis Category (%) |  |  |  |  |  |  |  |  |  |  |
| Asperger's Disorder | 23 ( 4.4%) |  | 16 ( 5.1%) |  | 4 ( 3.8%) |  | 3 ( 2.9%) |  | 92 ( 6.9%) |  |
| Autism or Autistic Disorder | 36 ( 6.9%) |  | 23 ( 7.3%) |  | 7 ( 6.7%) |  | 6 ( 5.8%) |  | 117 ( 8.8%) |  |
| Autism Spectrum Disorder | 438 (84.1%) |  | 258 (82.4%) |  | 89 ( 85.6%) |  | 91 ( 87.5%) |  | 1053 (79.2%) |  |
| Pervasive Developmental Disorder - Not Otherwise Specified (PDD-NOS) | 24 ( 4.6%) |  | 16 ( 5.1%) |  | 4 ( 3.8%) |  | 4 ( 3.8%) |  | 68 ( 5.1%) |  |
| History of Intellectual Disability (%) |  |  |  |  |  |  |  |  |  |  |
| Yes | 51 ( 9.8%) |  | 30 ( 9.6%) |  | 8 ( 7.7%) |  | 13 ( 12.5%) |  | 185 (14.5%) |  |
| No | 470 (90.2%) |  | 283 (90.4%) |  | 96 (92.3%) |  | 91 (87.5%) |  | 1161 (86.3%) |  |
| Functional language level (%) |  |  |  |  |  |  |  |  |  |  |
| Nonverbal | 16 ( 3.1%) |  | 10 ( 3.2%) |  | 4 ( 3.8%) |  | 2 ( 1.9%) |  | 164 (12.9%) |  |
| Single words | 36 ( 6.9%) |  | 24 ( 7.7%) |  | 6 ( 5.8%) |  | 6 ( 5.8%) |  | 182 (14.3%) |  |
| Phrase speech | 87 (16.7%) |  | 229 (73.2%) |  | 79 ( 76.0%) |  | 74 ( 71.2%) |  | 198 (15.5%) |  |
| Complex sentences | 382 (73.3%) |  | 50 (16.0%) |  | 15 ( 14.4%) |  | 22 ( 21.2%) |  | 731 (57.3%) |  |
| Full Scale IQ<sup>‡</sup> (mean (SD)) |  | 84.04 (22.85) |  | 84.10 (22.78) |  | 83.10 (21.88) |  | 84.80 (24.20) |  | 77.89 (23.38) |
| Nonverbal IQ (mean (SD)) |  | 94.87 (19.95) |  | 95.90 (20.09) |  | 93.30 (19.84) |  | 93.48 (19.74) |  | 91.75 (20.83) |
| Verbal IQ (mean (SD)) |  | 89.72 (21.30) |  | 89.96 (21.09) |  | 88.01 (20.94) |  | 90.80 (22.40) |  | 88.83 (22.52) |
| Missing rate among 271 features |  |  |  |  |  |  |  |  |  |  |
| No missing | 521 (100%) |  | 313 (100%) |  | 104 (100%) |  | 104 (100%) |  | 197 (14.6%) |  |
| <0.1 |  |  |  |  |  |  |  |  | 528 (39.2%) |  |
| 0.1-0.3 |  |  |  |  |  |  |  |  | 128 ( 9.5%) |  |
| 0.3-0.5 |  |  |  |  |  |  |  |  | 366 (27.2%) |  |
| 0.5-0.7 |  |  |  |  |  |  |  |  | 127 ( 9.4%) |  |

<sup>†</sup>SD: standard deviation
<sup>‡</sup>IQ: intelligence quotient

To compare the machine learning prediction performance with traditional methods, we applied the multiple imputation method to the same dataset of 521 participants and 271 measures. We kept the cognitive impairment status (FSIQ<80; FSIQ<70) in the training set available (n=313) and set the cognitive impairment status as missing in the initial testing set (n=104). We ran the multiple imputation algorithm (Van Buuren et al., 2011) to impute the missing cognitive impairment using the R package mice (Version 3.13.0). We assessed the accuracy, Cohen’s kappa, sensitivity, specificity and positive and negative predictive values (PPV, NPV) and compared the performance with the machine learning prediction.

### 6. Assess prediction performance in the additional independent dataset (n=1,346) with missingness

We used 1,346 newly collected participants with FSIQ, having no overlap with the original 521 cases and a missing rate of up to 70% among the 271 predictive features (**Table 1**). We first applied nonparametric missing value imputation methods (Stekhoven et al., 2012) to the dataset with all predictive features by R package missForest (Version 1.4). Then, we used our trained elastic net model to obtain the predicted probability of imputed cognitive impairment. We assessed the prediction performance by ROC curves with the predicted probability, and used the same probability cutoff as the initial testing set to assess the accuracy, Cohen’s kappa, sensitivity, specificity and positive and negative predictive values (PPV, NPV). We also assessed the prediction performance by different levels of missing rate among 271 features.

## Results

### 1. IQ scores in SPARK are highly linearly inter-correlated and independent of age

Among 521 ASD participants with complete FSIQ and all 271 measures (**Figure 1**) in the initial set, 81.2% were male (**Table 1**) and the average age at enrollment was 7.9 years (Range 2 – 15.5 years). The racial composition is 72.4% white, 7.1% African American, 1.7% Asian, 0.2% Native American, 1.3% other, and 17.3% more than one race. Mean IQ scores are: FSIQ=84.0, NVIQ=94.9, VIQ=89.7. The training, validation and testing sets have similar demographic distribution (**Table 1**).

Based on the scatterplots (**Supplementary Figure 1**) and the pair-wise Pearson correlation, FSIQ, NVIQ and VIQ are highly positively linearly correlated (r > 0.7). We also assessed the relationship between IQ scores and age of IQ testing (**Supplementary Figure 2**). There is no correlation between the three types of IQ scores and age (−0.06<r<0.06).

### 2. Machine learning models can accurately predict cognitive impairment as defined by FSIQ<80

We conducted an initial assessment of several measures considered to be relevant to IQ (**Table 2, Supplementary Figure 3**). Parent-reported severity of cognitive delay and functional language level are strongly associated with FSIQ, NVIQ and VIQ, followed by any language delay or language disorder, severity of language regression, and parent-reported ID diagnosis (**Table 2**).

**Table 2:**
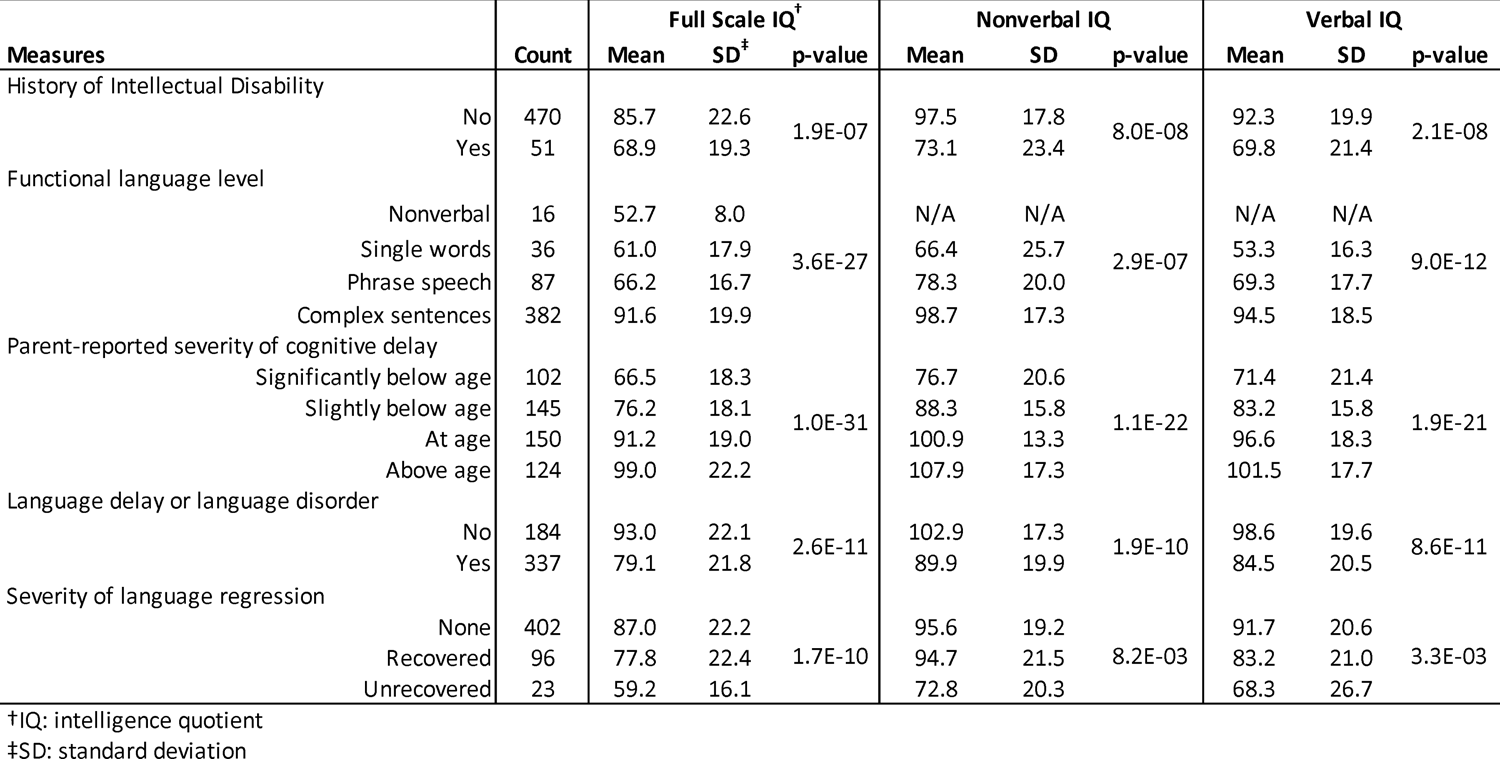
Association between cognitive, language measures and IQ scores

| Measures | Count | Full Scale IQ<sup>†</sup> |  |  | Nonverbal IQ |  |  | Verbal IQ |  |  |
| --- | --- | --- | --- | --- | --- | --- | --- | --- | --- | --- |
|  |  | Mean | SD<sup>‡</sup> | p-value | Mean | SD | p-value | Mean | SD | p-value |
| History of Intellectual Disability |  |  |  |  |  |  |  |  |  |  |
| No | 470 | 85.7 | 22.6 | 1.9E-07 | 97.5 | 17.8 | 8.0E-08 | 92.3 | 19.9 | 2.1E-08 |
| Yes | 51 | 68.9 | 19.3 |  | 73.1 | 23.4 |  | 69.8 | 21.4 |  |
| Functional language level |  |  |  |  |  |  |  |  |  |  |
| Nonverbal | 16 | 52.7 | 8.0 | 3.6E-27 | N/A | N/A | 2.9E-07 | N/A | N/A | 9.0E-12 |
| Single words | 36 | 61.0 | 17.9 |  | 66.4 | 25.7 |  | 53.3 | 16.3 |  |
| Phrase speech | 87 | 66.2 | 16.7 |  | 78.3 | 20.0 |  | 69.3 | 17.7 |  |
| Complex sentences | 382 | 91.6 | 19.9 |  | 98.7 | 17.3 |  | 94.5 | 18.5 |  |
| Parent-reported severity of cognitive delay |  |  |  |  |  |  |  |  |  |  |
| Significantly below age | 102 | 66.5 | 18.3 | 1.0E-31 | 76.7 | 20.6 | 1.1E-22 | 71.4 | 21.4 | 1.9E-21 |
| Slightly below age | 145 | 76.2 | 18.1 |  | 88.3 | 15.8 |  | 83.2 | 15.8 |  |
| At age | 150 | 91.2 | 19.0 |  | 100.9 | 13.3 |  | 96.6 | 18.3 |  |
| Above age | 124 | 99.0 | 22.2 |  | 107.9 | 17.3 |  | 101.5 | 17.7 |  |
| Language delay or language disorder |  |  |  |  |  |  |  |  |  |  |
| No | 184 | 93.0 | 22.1 | 2.6E-11 | 102.9 | 17.3 | 1.9E-10 | 98.6 | 19.6 | 8.6E-11 |
| Yes | 337 | 79.1 | 21.8 |  | 89.9 | 19.9 |  | 84.5 | 20.5 |  |
| Severity of language regression |  |  |  |  |  |  |  |  |  |  |
| None | 402 | 87.0 | 22.2 | 1.7E-10 | 95.6 | 19.2 | 8.2E-03 | 91.7 | 20.6 | 3.3E-03 |
| Recovered | 96 | 77.8 | 22.4 |  | 94.7 | 21.5 |  | 83.2 | 21.0 |  |
| Unrecovered | 23 | 59.2 | 16.1 |  | 72.8 | 20.3 |  | 68.3 | 26.7 |  |

†IQ: intelligence quotient
‡SD: standard deviation

Among 150 participants with FSIQ<70, 25 participants (16.7%) were reported as ID by their parents. With FSIQ<80, 37 out of 216 participants (17.1%) were reported as ID.

Machine learning models showed good prediction of cognitive impairment as defined by FSIQ<80. In the training set, we trained five commonly used machine learning models and assessed their prediction performance in the validation set by receiver operating characteristic (ROC) curves (**Figure 2**). Elastic-net regularized generalized linear model has the best performance (AUC=0.900) followed by support vector machines (AUC=0.891), k-nearest neighbors (AUC=0.863), random forest (AUC=0.863) and gradient boosting (AUC=0.843). Based on the prediction performance in the validation set, we chose the elastic-net model as the final model using 129 predictive features (**Supplementary table 1**).

**Figure 2:**
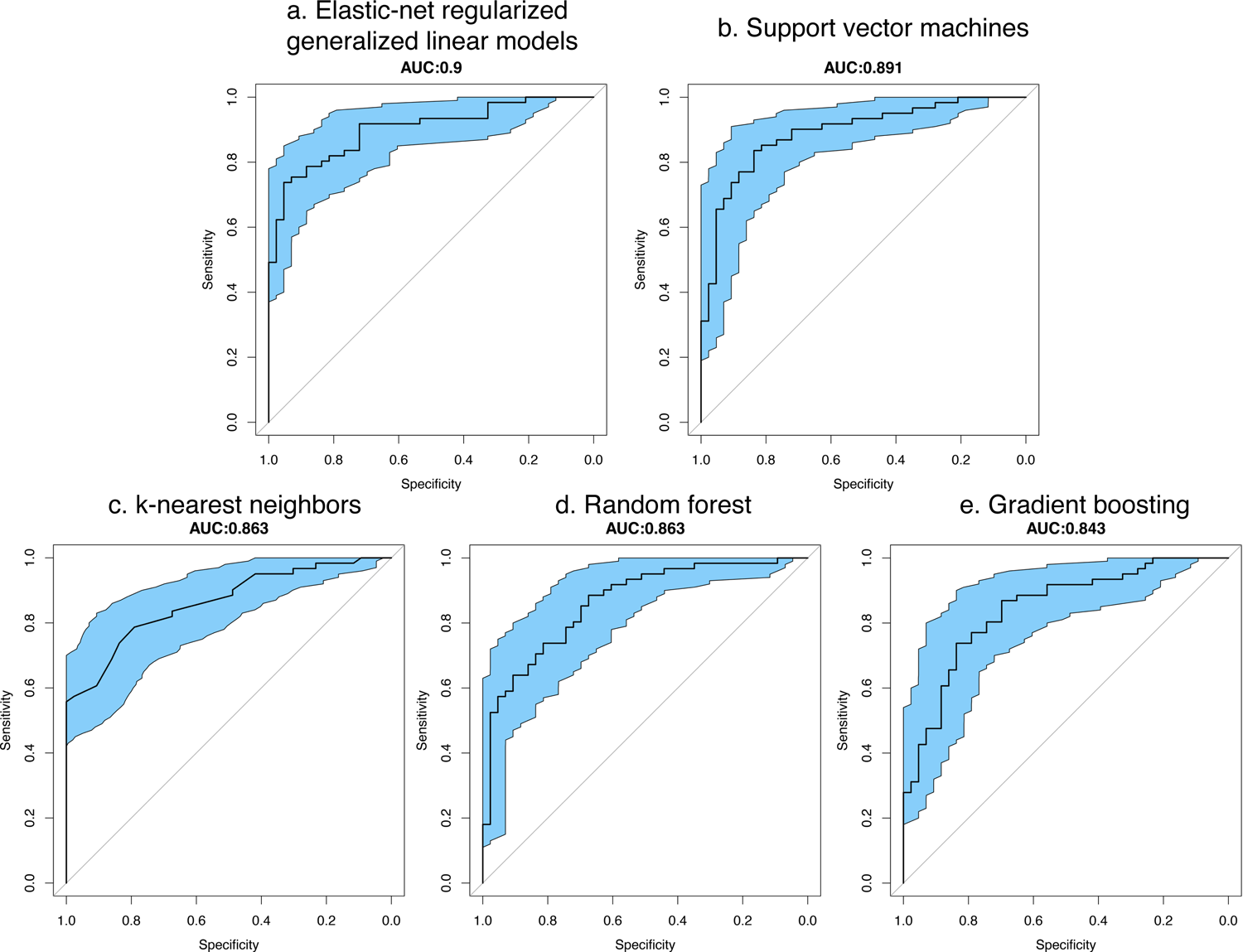
Prediction performance in validation sets among machine learning models for FSIQ<80 by receiver operating characteristic (ROC) curves

We calculated the variable importance with directionality for these 129 selected predictive features (**Figure 3, Supplementary table 1**). Parent-reported language and cognitive levels or diagnoses, age at ASD diagnosis, mood/anxiety disorder, specific items on standardized motor and ASD questionnaires and service history are significant IQ predictors.

**Figure 3:**
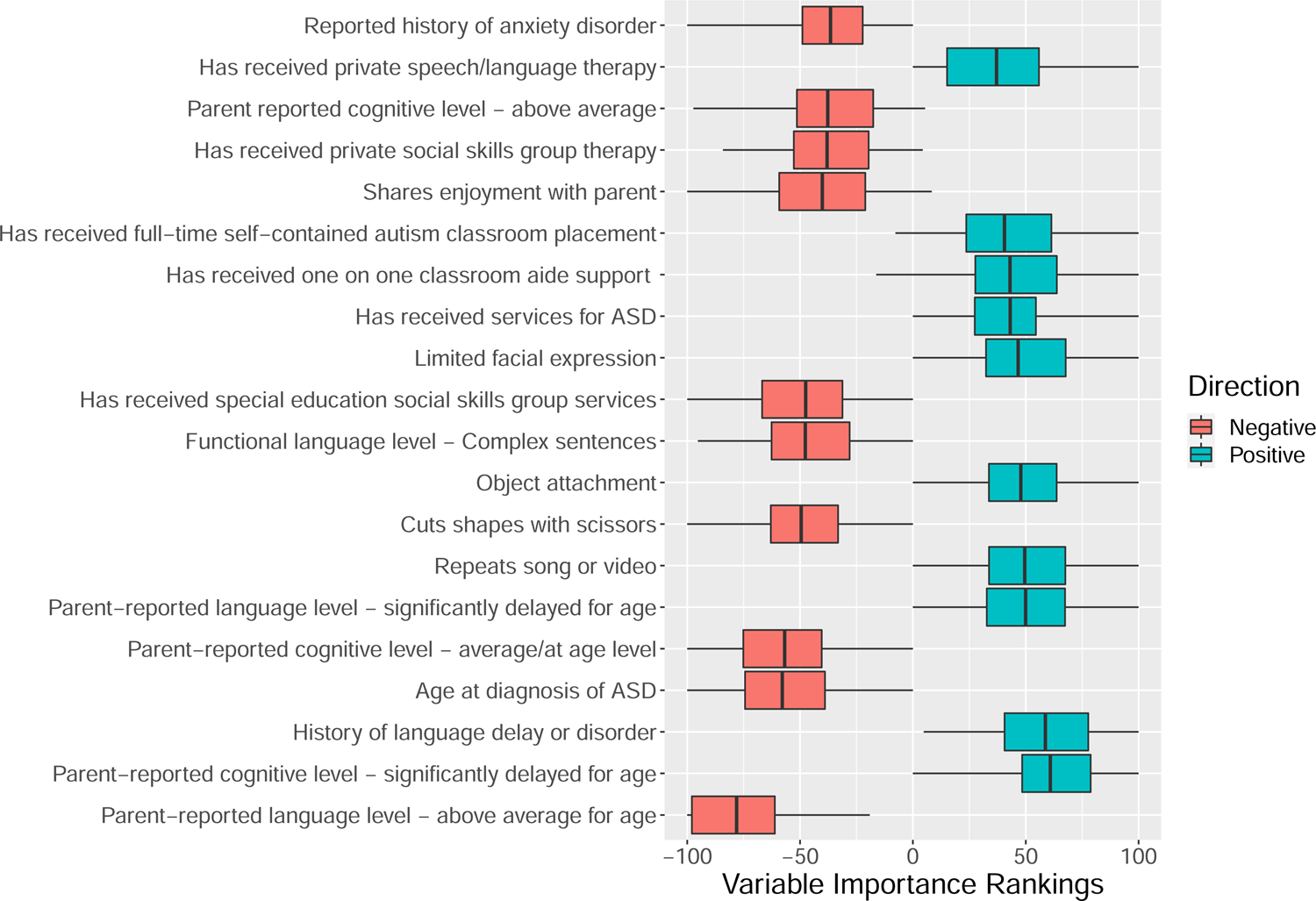
Top 20 variable importance rankings by elastic net model predicting FSIQ<80, scaled to −100 to 100. The absolute value of ranks reflects the relative contribution to the prediction of the model. Variables with positive ranks are positively associated with lower IQ, while variables with negative ranks are negatively associated with lower IQ

When applying our final elastic net model to the initial independent testing set, the predictive performance is relatively high with an AUC of 0.888 (**Figure 4**). With a cutoff of 0.45 on the predictive probability by the elastic-net model, the sensitivity is 0.744 and the specificity is 0.885 for predicting cognitive impairment (**Figure 4, Table 3**).

**Figure 4:**
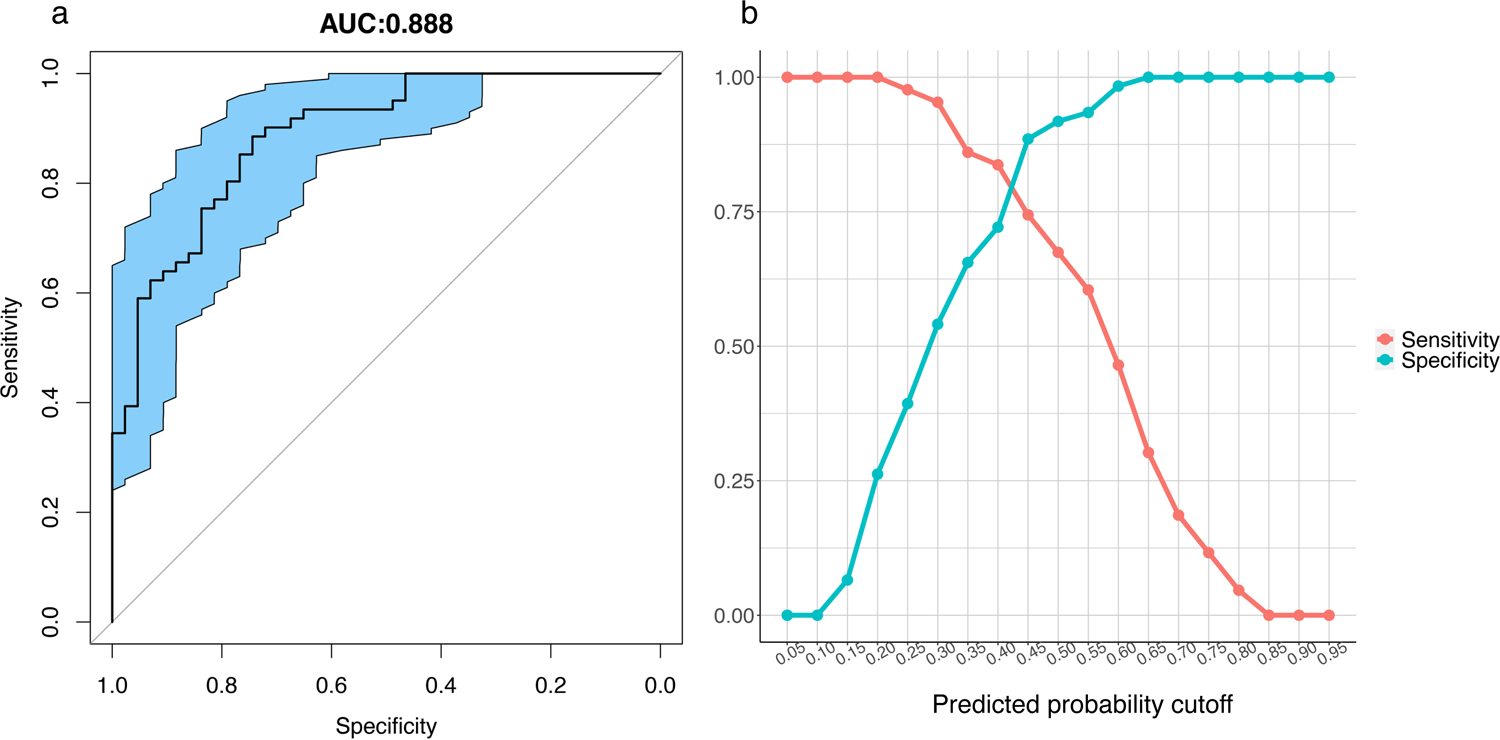
Prediction performance in initial testing set (n=104) by elastic-net model for FSIQ<80. a. Receiver operating characteristic (ROC) curves showing the area under the curve (AUC) is 0.888 in the testing set; b. Prediction probability cutoff by sensitivity and specificity. Final prediction probability cutoff of 0.45 would provide a sensitivity of 0.744 and a specificity of 0.885

**Table 3:** Prediction performance by elastic net classification models and multiple imputation

|  | Elastic net classification model |  |  |  | Multiple imputation |  |
| --- | --- | --- | --- | --- | --- | --- |
| FSIQ<sup>†</sup> Cutoff | FSIQ<80 |  | FSIQ<70 |  | FSIQ<80 | FSIQ<70 |
| Dataset | Initial testing set (n=104) | Additional independent set (n=1,346) | Initial testing set (n=104) | Additional independent set (n=1,346) | Initial testing set (n=104) | Initial testing set (n=104) |
| AUC<sup>‡</sup> | 0.888 | 0.876 | 0.881 | 0.865 | N/A | N/A |
| Sensitivity | 0.744 | 0.772 | 0.767 | 0.750 | 0.558 | 0.600 |
| Specificity | 0.885 | 0.803 | 0.838 | 0.830 | 0.787 | 0.851 |
| Positive Predictive Value | 0.821 | 0.805 | 0.657 | 0.744 | 0.649 | 0.621 |
| Negative Predictive Value | 0.831 | 0.769 | 0.899 | 0.834 | 0.716 | 0.840 |
| Accuracy | 0.827 | 0.787 | 0.817 | 0.798 | 0.692 | 0.779 |
| Kappa | 0.638 | 0.574 | 0.576 | 0.579 | 0.352 | 0.456 |

†FSIQ: Full scale intelligence quotient
‡AUC: area under the curve

We compared the model’s performance with an multiple imputation approach (**Table 3**). The overall accuracy, kappa, sensitivity and specificity are lower by multiple imputation compared to the elastic net model (**Table 3**; FSIQ<80: accuracy=0.692, sensitivity=0.558, specificity=0.787; FSIQ<70: accuracy=0.779, sensitivity=0.600, specificity=0.851).

### 3. Prediction performance for FSIQ<80 in an additional independent set (n=1,346) in SPARK with missingness in predictive features

In the larger newly collected independent set with missing rate up to 70% (n=1,346; **Figure 1, Table 1**):12.9% are nonverbal, 14.5% reported ID and the mean FSIQ is 77.89 (**Table 1**). Among all 271 features, rates of each level of missing values in the sample are as follows: no missing −197 (14.6%), <10% missing-528 (39.2%), 10-30% missing - 128 (9.5%), 30-50% missing - 366 (27.2%) and 50-70% missing - 127 (9.4%) participants (**Table 1**).

We imputed the missing values among predictive features by existing methods (Stekhoven et al., 2012) and then applied our trained elastic net model to obtain predicted probability of imputed cognitive impairment (FSIQ<80). By using a cutoff of 0.45 on the predicted probability, most prediction performance metrics in the additional independent set are comparable and just slightly lower than in the initial testing set (AUC=0.876, **Figure 5**; sensitivity=0.772, specificity=0.803, accuracy=0.787, **Table 3**). We also examined the prediction performance in subsets of participants with different missing rates within the additional independent set. The AUC’s among all subsets were comparable to the AUC in the initial testing set with no missing values (n=104) (**Supplementary Figure 6a, Supplementary Table 4**). Overall, the prediction performance is still comparable to the initial testing set even with some level of missingness among the predictors.

### 4. Sensitivity analysis when cognitive impairment is defined by FSIQ<70

The elastic net model for FSIQ<70 selected 128 features, and top ranked predictors are similar to the previous model predicting FSIQ<80. In both the initial testing set (n=104) and the additional independent set (n=1,346), most prediction performance metrics in the FSIQ<70 model are comparable and just slightly lower than in the FSIQ<80 model (initial testing set: AUC=0.881; additional independent set: AUC=0.865, **Table 3, Supplementary Figure 4b**). The prediction performance between the initial testing set and the additional independent set is comparable.

**Figure 5:**
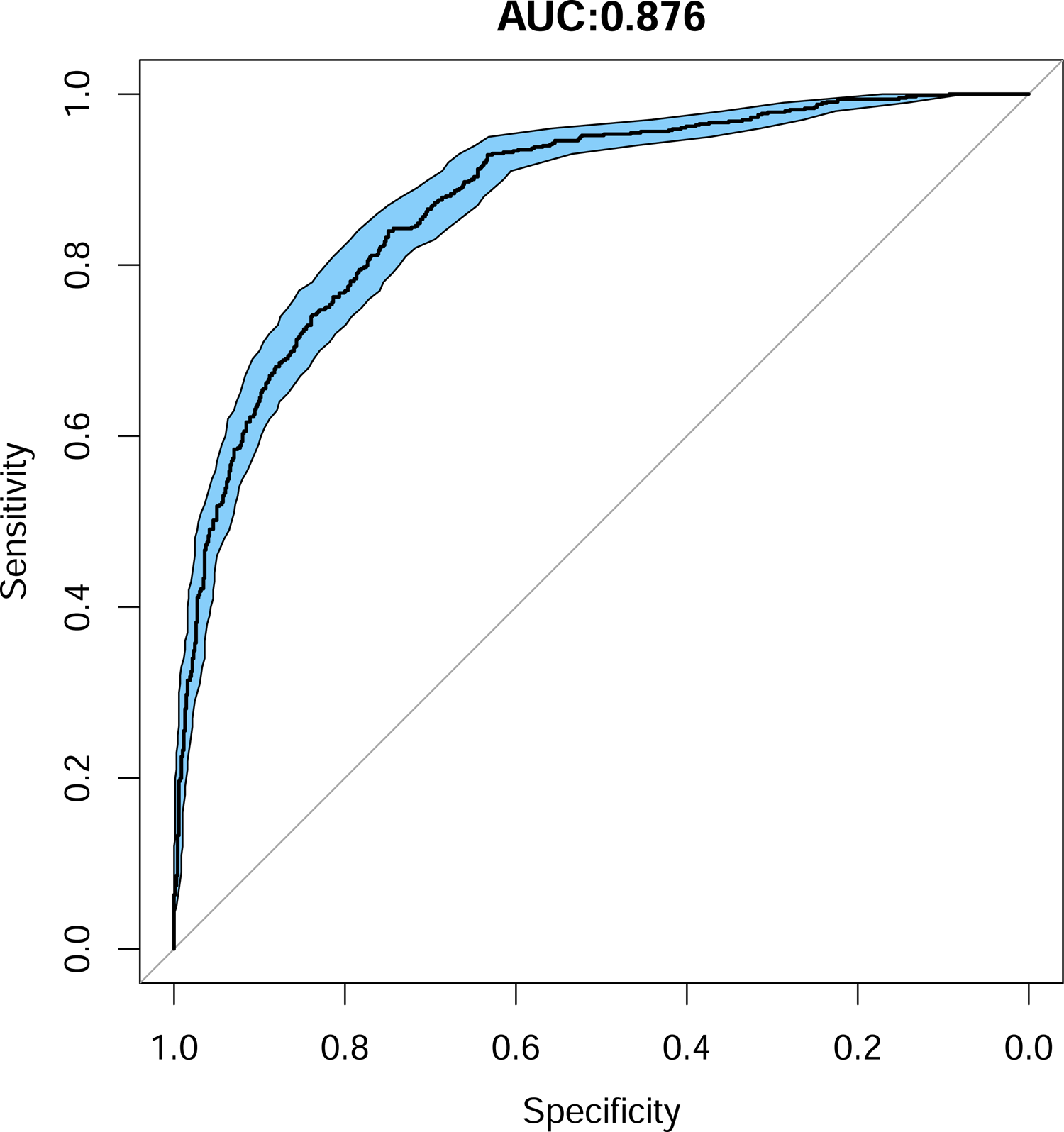
Prediction performance in the additional independent set (n=1,346) by elastic-net model for FSIQ<80. Receiver operating characteristic (ROC) curves showing the area under the curve (AUC) is 0.876.

### 5. Predicting continuous FSIQ scores by machine learning methods

We trained the elastic net, support vector machines and k-nearest neighbors models to predict continuous FSIQ, and evaluated their performances compared to the binary prediction models in the initial testing set (n=104) and additional independent set (n=1,346; **Supplementary Table 3, Supplementary figure 5**). The elastic net model has relatively good prediction performance and similar variables of importance. The Pearson correlation with true FSIQ is 0.678 in the initial testing set (n=104) and 0.730 in the additional independent set (n=1,346), while the root mean square error (RMSE) is 17.4 in the initial testing set and 16.1 in the additional independent set (**Supplementary Table 3**). We also examined the RMSE in subsets of participants with different missing rate in the additional independent set (n=1,346): the RMSE among all subsets was comparable to the initial testing set with no missing values (n=104) (**Supplementary Figure 6c, Supplementary Table 4**). The support vector machines model also has similar prediction performance to the elastic net model in predicting continuous FSIQ (**Supplementary Table 3,4**).

We then compared the prediction performance of the continuous FSIQ model to our binary cognitive impairment model, using binary evaluation metrics. To do this, we categorized predicted continuous FSIQ according to cutoffs (<80 and <70, respectively) and compared it to true FSIQ classified by the same cutoffs (**Supplementary Table 3**). Overall, compared to the original binary elastic net prediction model, the prediction performance was comparable (initial testing set: FSIQ<80 AUC=0.864, FSIQ<70 AUC=0.812; additional independent set: FSIQ<80 AUC=0.871, FSIQ<70 AUC=0.875; **Supplementary Table 3, Table 3**). The prediction performance is also similar across subsets with varying missing rates in the additional independent set (**Supplementary Table 4**).

Because our main purpose in our research is to impute the presence or absence of cognitive impairment, we used the binary elastic net prediction as our final model.

## Discussion

Using machine learning methods, we demonstrate it is possible to derive valid estimates of cognitive ability based on parent-reported data in a large research cohort of autistic individuals for whom formal IQ testing is not available. We are also able to show our model performance is relatively stable in the presence of missing data and our approach is generalizable to impute cognitive impairment in the broader population of children with ASD. Having a cognitive estimate permits more sophisticated phenotypic and genomic analyses with available registry data, matching of appropriate control groups, and stratification or covariation in analyses for subtyping or to control for influences beyond core autism. It also fills a critical gap to include more underserved populations and more severely affected individuals for whom cognitive testing may not be available.

We obtained the variable importance rankings by elastic-net model for all available measures in SPARK. Although the variable importance rankings are model dependent and data dependent, we found several interesting items that contribute to the prediction model. The assessment of predictive importance among all available measures in SPARK revealed that parent report on both standardized and non-standardized instruments contains valuable information regarding individuals’ cognitive level. Studies of other clinical conditions have included similar information as predictors of future cognitive status with good success, for example, demographic data and health history (Graham et al., 2020). In the present study, the strongest predictors were parent report of history of language delay and disorder, report of general cognitive delay, and younger age at ASD diagnosis.

We chose to focus on detection of IQ under 80, so as to sensitively capture borderline intellectual impairment for our research, beyond a more rigid and outdated definition of intellectual disability. We compared definitions of cognitive impairment as FSIQ under 80 or 70, and predictions using either definition are accurate, suggesting generalizability. However, surprisingly few parents reported ID in children with a tested IQ below 70 or 80. Parent-report of ID diagnosis was not as strongly associated with IQ scores and not as successful as other parent reported information in predicting lower IQ. These findings are consistent with prior studies showing parents tend to underreport developmental disabilities (Porter et al., 2011). We found that more concrete questions about day-to-day observations of language and cognitive tasks relative to age are more significantly associated with IQ scores and are more accurate predictors of cognitive impairment. The predictive importance of parent-reported ID diagnosis was low, and lower than many other reported characteristics, including autistic behaviors, early social-communication skills, and psychiatric co-morbidities. One of the most interesting autism characteristics that served to predict cognitive impairment was repetitive playing of a song or video segment. In terms of observable skills, after language and overall reasoning ability, good fine motor control with scissors was an indicator that cognitive impairment is unlikely. Other strong components were education service history: receipt of specific autism services, a one-on-one classroom aide and/or assignment to a full time self-contained ASD classroom all indicate likely cognitive impairment, while assignment to social skills group therapy at school indicates the likelihood that IQ is within normal limits.

There were many low-ranking variables with minimal to no predictive importance in the model that are notable. For example, a history of generic special education or early intervention, and several early signs (such as speech delay and absence of reciprocal smiling) had low yield. Similarly, age, maternal higher education, overall severity of repetitive behavior or number of signs of autism, sleep problems, motor delays, and history of a non-language regression were not strong predictors. On the standardized measures, while some specific repetitive and basic social behaviors are predictive, others are not. The reason these factors failed to be selected by the model is unclear: it is possible some are universal features of autism independent of cognitive ability, had low base rates in our sample, or were highly correlated with other variables that were alternatively selected by the model.

There are several potential implications of our findings. First, when surveying cognitive ability in large cohorts, it is important to anchor against chronological age expectations and to be specific about the skills queried. IQ and mental age data requested from families are often missing. Second, consistent with prior literature, language level remains the strongest correlate of IQ. Finally, the present model suggests a set of variables that can be easily administered as survey questions and used in an algorithm to predict cognitive ability for children in autism studies.

There are several limitations to the present study. First, the current model included only children under 18, and results could be different for adults. Second, the assumptions of the model require certain caveats. Not all individuals with poor past performance on a single IQ test have true global cognitive impairment or ID, nor do all individuals with limited language. Individuals must be appreciated for their abilities rather than their “disabilities,” and this is the purpose of expert, comprehensive, in-person assessment. In this way, all statistical models risk inaccuracy when applied to an individual. Caution is urged in estimating cognitive impairment in children below school age in particular.

The current model was trained on IQ testing data completed at any age or point in time; however, if planning to test this or a similar model, researchers may wish to exclude cases if IQ tests were completed a long time ago or prior to age 5.

The algorithm for imputing cognitive level may have more limited generalizability to groups outside of SPARK given the relatively high IQ and basic language ability of participants in the final dataset relative to population estimates in autism (Baio et al., 2018), and the high SES reported previously in SPARK (SPARK Consortium, 2017). The problem of discrepancies between NVIQ and VIQ in many children with autism also must be considered. Although NVIQ correlated highly with FSIQ in our sample, there is greater variability in its association with VIQ, and our analysis necessarily excluded children who had no FSIQ but only a NVIQ or derived ratio IQ available and thus may have been nonverbal. In applying an algorithm that leans quite heavily on language variables and is based upon FSIQ as the outcome, researchers must remain aware of the possibility the model could underestimate some nonverbal individuals. However, despite these concerns, it is promising that accuracy of the model for the subset of nonverbal individuals in our testing set was high (accuracy=97% among 164 non-verbal participants).

When interpreting the variables of greatest importance in the model, it is important to note the concept of importance is model-dependent and data dependent, and each characteristic on its own cannot be assumed as a one-to-one proxy for IQ classification or ID in an individual, but rather operate together to maximize predictive power. Further, adaptive behavior, the current standard for ID diagnosis, was not included in the analysis due to missing data. Regardless, taken together, the variables clearly have face validity and clinical relevance given our knowledge of children with greater impairments (Bishop et al., 2015; Munson et al., 2008).

In terms of methodology, despite our use of regularization and cross-validation to minimize overfitting, our ratio of data-to-features was relatively low in our model development, which could lead to overfitting and limit the model’s performance when imputing cognitive level in other ASD populations. However, we showed the model’s performance was comparably good in an additional independent set even in the presence of missing data.

Future work will re-train and replicate the predictive models with larger samples providing a greater data-to-features ratio and more complete cases with additional phenotyping data, and will evaluate the reliability of medical record data abstraction underlying the IQ data. Dissecting ASD subtypes by intellectual functioning level will allow us to more fully stratify and thereby understand similarities and differences in underlying biology across the autism spectrum. The model stands to provide a critical missing piece for future genomic and other analyses in the SPARK cohort and beyond.

---

**Supplementary Figure 1:** Full Scale IQ, Nonverbal IQ and Verbal IQ are linearly correlated

**Supplementary Figure 2:** Full Scale IQ, Nonverbal IQ and Verbal IQ are independent of age

**Supplementary Figure 3:** Full Scale IQ by cognitive and language measures by boxplot

**Supplementary Figure 4:** Prediction performance in initial testing set by elastic-net model for FSIQ<70. a. receiver operating characteristic (ROC) curves showing the area under the curve (AUC) is 0.881 in the testing set; b. Prediction probability cutoff by sensitivity and specificity. Final cutoff of 0.35 would provide a sensitivity of 0.767 and a specificity of 0.838

**Supplementary Figure 5:** Predicted FSIQ versus true FSIQ by elastic-net, support vector machines (SVM) and k-nearest neighbors (kNN) in the initial testing set (n=104) and in the additional independent set (n=1,346)

**Supplementary Figure 6:** Prediction performance by missing rate in the additional independent set (n=1,346). Panels a, b panel show the AUC by missing rate based on the elastic-net model trained to predict cognitive impairment defined by FSIQ<80 (a) and FSIQ<70 (b). The grey dashed line is the AUC from the testing set in the model development (AUC=0.888 for FSIQ<0.8 and AUC=0.881 for FSIQ<0.7). Panel c shows the root mean square error (RMSE) by missing rate in the additional independent set compared to the testing set in model development, for three models predicting continuous FSIQ

## Conflict of Interest

All authors declare that there is no conflict of interest.

## Data Availability

All data for the SPARK population dataset described in this study are available to researchers in the Simons Foundation Autism Research Initiative database, SFARI Base, at https://base.sfari.org.

## Acknowledgements

We are grateful to all of the families in SPARK, the SPARK clinical sites and SPARK staff. We appreciate obtaining access to the SPARK phenotypic dataset on SFARI Base. Approved researchers can obtain the SPARK population dataset described in this study by applying at https://base.sfari.org. The SPARK consortium members at the time of this writing are: Leonard Abbeduto, Gabriella Aberbach, Shelley Aberle, John Acampado, Andy Ace, Kaitlyn Ahlers, Charles Albright, Michael Alessandri, Nicolas Alvarez, David Amaral, Alpha Amatya, Alicia Andrus, Claudine Anglo, Rob Annett, Eduardo Arzate, Irina Astrovskaya, Kelli Baalman, Melissa Baer, Gabriele Baraghoshi, Nicole Bardett, Sarah Barnes, Asif Bashar, Heidi Bates, Katie Beard, Juana Becerra, Malia Beckwith, Landon Beeson, Josh Beeson, Brandi Bell, Monica Belli, Dawn Bentley, Natalie Berger, Anna Berman, Raphael Bernier, Elizabeth BerryKravis, Mary Berwanger, Shelby Birdwell, Elizabeth Blank, Stephanie Booker, Aniela Bordofsky, Erin Bower, Catherine Bradley, Stephanie Brewster, Elizabeth Brooks, Aliso Brown, Melissa Brown, Jennylyn Brown, Cate Buescher, Martin Butler, Eric Butter, Wenteng CaI, Norma Calderon, Kristen Callahan, Alexies Camba, Claudia CampoSoria, Paul Carbone, Laura Carpenter, S. Carpenter, Lindsey Cartner, Myriam Casseus, Lucas Casten, Sullivan Catherine, Ashley Chappo, Tia Chen, Wubin Chin, Sharmista Chintalapalli, Daniel Cho, Dave Cho, YB Choi, Wendy Chung, Renee Clark, Cheryl Cohen, Kendra Coleman, Costanza Colombi, Joaquin Comitre, Sarah Conyers, Lindsey Cooper, Leigh Coppola, Lisa Cordiero, Jeanette Cordova, Dahriana Correa, Hannah Cottrell, Michelle Coughlin, Eric Courchesne, Dan Coury, Joseph Cubeis, Sean Cunningham, Mary Currin, Michele Cutri, Sophia D’Ambrosi, Amy Daniels, Sabrina Davis, Nickelle Decius, Jennifer Delaporte, Brandy Dennis, Kate Dent, Gabrielle Dichter, Katharine Diehl, Chris Diggins, Emily Dillon, Erin Doyle, Andrea Drayton, Megan DuBois, Gabrielle Duhon, Megan Dunlevy, Rachel Earl, Catherine Edmonson, Sara Eldred, Barbara Enright, Craig Erickson, Amy Esler, Anne Fanta, Carrie Fassler, Faris Fazal, Pam Feliciano, Angela Fish, Kate Fitzgerald, Chris Fleisch, Eric Fombonne, Emily Fox, Sunday Francis, Margot Frayne, Sandra Friedman, Laura Fuller, Virginia Galbraith, Swami Ganesan, Jennifer Gerdts, Mohammad Ghaziuddin, Haidar Ghina, David Giancarla, Erin Given, Jared Gong, Kelsey Gonring, Natalia Gonzalez, Antonio Gonzalez, Rachel Gordon, Catherine Greay, LeeAnne Green Snyder, Tunisia Greene, Ellen Grimes, Luke Grosvenor, Amanda Gulsrud, Abha Gupta, Jaclyn Gunderson, Chris Gunter, Anibal Gutierrez, Frampton Gwynette, Melissa Hale, Lauren K. Hall, Jake Hall, Kira Hamer, Bing Han, Nathan Hanna, Antonio Hardan, Eldric Harrell, Jill Harris, Nina Harris, Caitlin Hayes, Teryn Heckers, Kathryn Heerwagen, Susan Hepburn, Lynette Herbert, Clara Herrera, Brittani Hilscher, Kathy Hirst, Theodore Ho, Dabney Hofammann, Margaret Hojlo, Gregory Hooks, Dain Howes, Lark Huang-Storm, Samantha Hunter, Hanna Hutter, Teresa Ibanez, Dalia Istephanous, Suma Jacob, Andrea Jarratt, Stanley Jean, Anna Jelinek, Bill Jensen, Mya Jones, Mark Jones, Alissa Jorgenson, Roger Jou, Jessyca Judge, Taylor Kalmus, Stephen Kanne, Hannah Kaplan, Lauren Kasperson, Sophy Kim, Annes Kim, Cheryl Klaiman, Robin Kochel, Misia Kowanda, Melinda Koza, Sydney Kramer, Eva KurtzNelson, Hoa Lam, Elena Lamarche, Erica Lampert, Rebecca Landa, Alex Lash, Noah Lawson, J. Kiely Law, Holly Lechniak, CD Lehman, Bruce Leight, Laurie Lesher, Deana Li, Robin Libove, Natasha Lillie, Danica Limon, Desi Limpoco, Nathan Lo, Brandon Lobisi, Marilyn Lopez, Catherine Lord, Daniella Lucio, Addie Luo, Audrey Lyon, Natalie Madi, Malcolm Mallardi, Lacy Malloch, Anup Mankar, Lori Mann, Patricia Manning, Julie Manoharan, Olena Marchenko, Richard Marini, Christa Martin, Gabriela Marzano, Sarah Mastel, Sheena Mathai, Clara Maxim, Caitlin McCarthy, Nicole Mccoy, Julie McGalliard, Anne-Marie McIntyre, Brooke McKenna, Alexander McKenzie, Megan McTaggart, Sophia Melnyk, Alexandra Miceli, Sarah Michaels, Jacob Michaelson, Anna Milliken, Amanda Moftt Gunn, Sarah Mohiuddin, Jessie Montezuma, Amy Morales-Lara, Kelly Morgan, Hadley Morotti, Michael Morrier, Maria Munoz, Karla Murillo, Kailey Murray, Vincent Myers, Natalie Nagpal, Jason Neely, Katelyn Neely, Olivia Newman, Richard Nguyen, Victoria Nguyen, Amy Nicholson, Melanie Niederhauser, Megan Norris, Kaela O’Brien, Eirene O’Connor, Mitchell O’Meara, Molly O’Neil, Brian O’Roak, Edith Ocampo, Cesar Ochoa-Lubinof, Jessica Orobio, Elizabeth Orrick, Crissy Ortiz, Opal Ousley, Motunrayo Oyeyemi, Samiza Palmer, Katrina Pama, Juhi Pandey, Katherine Pawlowski, Micah Pepper, Diamond Phillips, Karen Pierce, Joseph Piven, Jose Polanco, Natalie Pott-Schmidt, Lisa Prock, Angela Rachubinski, Desiree Rambeck, Rishiraj Rana, Shelley Randall, Vaikunt Ranganathan, Ashley Raven, Madelyn Rayos, Kelli Real, Louis F. Reichardt, Richard Remington, Anna Rhea, Catherine Rice, Harper Richardson, Stacy Rife, Chris Rigby, Ben Right, Beverly Robertson, Erin Roby, Casey Roche, Nicki Rodriguez, Katherine Roeder, Daniela Rojas, Cordelia Rosenberg, Jacob Rosewater, Katelyn Rossow, Payton Runyan, Nicole Russo, Tara Rutter, Mahfuza Sabiha, Mustafa Sahin, Marina Sarris, Dustin Sarver, Madeline Savage, Jessica Scherr, Hayley Schools, Gregory Schoonover, Robert Schultz, Brady Schwind, Cheyanne Sebolt, Rebecca Shafer, Swapnil Shah, Neelay Shah, Roman Shikov, Mojeeb Shir, Amanda Shocklee, Clara Shrier, Lisa Shulman, Matt Siegel, Andrea Simon, Laura Simon, Kaitlyn Singer, Emily Singer, Vini Singh, Kaitlin Smith, Chris Smith, Ashlyn Smith, Latha Soorya, John Spiro, Diksha Srishyla, Danielle Stamps, Laura Stchur, Morgan Steele, Alexandra Stephens, Amy Swanson, Megan Sweeney, Anthony Sziklay, Maira Tafolla, Nicole Takahashi, Amber Tallbull, Nicole Targalia, Cora Taylor, Sydney Terroso, Angela Tesng, Samantha Thompson, Jennifer Tjernagel, Jaimie Toroney, Laina Townsend, Katherine Tsai, Ivy Tso, Maria Valicenti-Mcdermott, Bonnie VanMetre, Candace VanWade, Dennis Vasquez Montes, Alison Vehorn, Mary Verdi, Brianna Vernoia, Natalia Volfovsky, Lakshmi Vrittamani, Jermel Wallace, Corrie Walston, Audrey Ward, Zachary Warren, William Curtis Weaver, Sabrina White, L. Casey White-Lehman, Fiona Winoto, Ericka Wodka, Jessica Wright, Sabrina Xiao, Simon Xu, WhaJames Yang, Amy Yang, Meredith Yinger, Christopher Zaro, Hana Zaydens, Cindy Zha, Allyson Zick

## Notes

### Competing Interest Statement

The authors have declared no competing interest.

### Funding Statement

This work is supported by the Simons Foundations

### Author Declarations

All protocols are approved by Western IRB, protocol #20151664.

## References

1. Allegrini, A. G., Selzam, S., Rimfeld, K. et al. (2019). Genomic prediction of cognitive traits in childhood and adolescence. Mol Psychiatry, 24, 819–827. https://doi.org/10.1038/s41380-019-0394-4.

2. Alvares, G. A., Bebbington, K., Cleary, D., Evans, K., Glasson, E. J., Mayberry, M. T., Pillar, S., Uljarevik, U., Varcin, K., Wray, J., Whitehouse A. J. O. (2020). The misnomer of ‘high functioning autism’: Intelligence is an imprecise predictor of functional abilities at diagnosis. Autism, 24, (1): 221–232.

3. Anderson, D. K., Liang, J. W., & Lord, C. (2014). Predicting young adult outcomes among more and less cognitively able individuals with autism spectrum disorders. Journal of Child Psychology and Psychiatry, 55(5), 485–494.

4. Baio, J., Wiggins L., Christensen D. L., et al. (2018). Prevalence of Autism Spectrum Disorder Among Children Aged 8 Years — Autism and Developmental Disabilities Monitoring Network, 11 Sites, United States, 2014. MMWR Surveill Summ 2018, 67(*SS-6)*, 1–23. http://dx.doi.org/10.15585/mmwr.ss6706a1externalicon.

5. Bayley, N., Aylward, G.P. (2019). Bayley Scales of Infant and Toddler Development. San Antonio, TX: Pearson.

6. Bishop, S. L., Farmer, C., Thurm, A. (2015). Measurement of Nonverbal IQ in Autism Spectrum Disorder: Scores in Young Adulthood compared to Early Childhood. Journal of Autism and Developmental Disorders, 45(*4*), 966–974.

7. Bodfish, J. W., Symons, F. J., Parker, D. E., & Lewis, M. H. (2000). Varieties of repetitive behavior in autism: comparisons to mental retardation. Journal of Autism and Developmental Disorders, 30(*3*), 237–243. doi: 10.1023/a:1005596502855

8. Bölte, S., Poustka, F. (2002). The Relation Between General Cognitive Level and Adaptive Behavior Domains in Individuals with Autism with and Without Co-Morbid Mental Retardation. Child Psychiatry Hum Dev, 33, 165–172. https://doi.org/10.1023/A:1020734325815

9. Chandler, S., Howlin, P., Simonoff, E., Kennedy, J., Baird, G. (2016). Comparison of parental estimate of developmental age with measured IQ in children with neurodevelopmental disorders. Child, 42(*4*), 486–493.

10. Charman, T., Pickles, A., Simonoff, E., Chandler, S., Loucas, T., & Baird, G. (2011). IQ in children with autism spectrum disorders: data from the Special Needs and Autism Project (SNAP). Psychological medicine, 41(*3*), 619–627. https://doi.org/10.1017/S0033291710000991

11. Chen, T., & Guestrin, C. (2016, August). Xgboost: A scalable tree boosting system. In Proceedings of the 22nd acm sigkdd international conference on knowledge discovery and data mining (pp. 785-794).

12. Coleman, K. J., Lutsky, M. A., Yau, V., et al. (2015). Validation of Autism Spectrum Disorder Diagnoses in Large Healthcare Systems with Electronic Medical Records. Journal of Autism and Developmental Disorders, 45(*7*),1989–96. 10.1007/s10803-015-2358-0.

13. Cornish, R. P., Tilling, K., Boyd, A., Davies, A., Macleod, J. (2015). Using linked educational attainment data to reduce bias due to missing outcome data in estimates of the association between the duration of breastfeeding and IQ at 15 years. Int J Epidemiol, 44(*3*), 937–45. 10.1093/ije/dyv035.

14. Corona, L. L., Wagner, L., Wade, J., Weitlauf, A. S., Hine, J., Nicholson, A., Stone, C., Vehorn, A., & Warren, Z. (2021). Toward Novel Tools for Autism Identification: Fusing Computational and Clinical Expertise. Journal of autism and developmental disorders, 1–10. Advance online publication. https://doi.org/10.1007/s10803-020-04857-x.

15. Dawson, M., Soulières, I., Gernsbacher, M. A., Mottron, L. (2007). The level and nature of autistic intelligence. Psychological Science, 18(*8*), 657–662.

16. De Rubeis, S., He, X., Goldberg, A., Poultney, C. S., Samocha, K., Cicek, A. E., et al. (2014). Synaptic, transcriptional and chromatin genes disrupted in autism. Nature, 515(*7526*), 209.

17. Elliott, C. D. (2007). Differential ability scales (2nd ed.). San Antonio: Harcourt Assessment.

18. Emerson, E., Hatton, C., Baines, S., & Robertson, J. (2016). The physical health of British adults with intellectual disability: cross sectional study. International journal for equity in health, 15(*1*), 1–9.

19. Feliciano, P., Zhou, X., Astrovskaya, I., Turner, T.N., Wang, T., Brueggeman, L., Barnard, R., Hsieh, A., Snyder, L. G., Muzny, D. M, et al. (2019). Exome sequencing of 457 autism families recruited online provides evidence for autism risk genes. NPJ Genome Med, 4(19).

20. Finch, W. H., Chang, M., Davis, A. S., Holden, J. E., Rothlisberg, B. A., & McIntosh, D. E. (2011). The prediction of intelligence in preschool children using alternative models to regression. Behavior research methods, 43(4), 942–952.

21. Fombonne, E., Coppola, L., Mastel, S., O’Roak, B. (2021). Validation of Autism Diagnosis and Clinical Data in the SPARK Cohort. Journal of Autism and Developmental Disorders. 10.1007/s10803-021-05218-y. Advance online publication. https://doi.org/10.1007/s10803-021-05218-y

22. Frazier, T. W., Demaree, H., Youngstrom, E. A. (2004). Meta-Analysis of Intellectual and Neuropsychological Test Performance in Attention-Deficit/Hyperactivity Disorder. Neuropsychology, 18(*3*), 543–555. DOI: 10.1037/0894-4105.18.3.543

23. Freeman, B. J., Ritvo, E. R., Yokota, A,, Childs, J., Pollard, J. (1998). WISC-R and Vineland Adaptive Behavior Scale Scores in Autistic Children. Journal of the American Academy of Child & Adolescent Psychiatry, 27(*4*), 428–429. DOI: 10.1038/s41525-019-0093-8.

24. Friedman, J., Hastie, T., & Tibshirani, R. (2001). The elements of statistical learning (Vol. 1, No. 10). New York: Springer series in statistics.

25. Graham, S. A, Lee, E. L., Jeste, D. V., Van Patten, R., Twamley, E. W., Nebeker, C., Yamadag, Y., Kim, H. C., Depp, C.A (2020). Artificial intelligence approaches to predicting and detecting cognitive decline in older adults: a conceptual review. Psychiatry Research, 284, 1–15.

26. Grove, J., Ripke, S., Als, T.D. et al. (2019). Identification of common genetic risk variants for autism spectrum disorder. Nat Genet 51, 431–444. https://doi.org/10.1038/s41588-019-0344-8

27. *Grzadzinski*, *R.*, Huerta, M., Lord, C. (2013). DSM-5 and autism spectrum disorders (ASDs): an opportunity for identifying ASD subtypes. Mol Autism, 4(*1*), 12.

28. Hamburg, S., Lowe, B., Startin, C. M., Padilla, C., et al. (2019). Assessing general cognitive and adaptive abilities in adults with Down syndrome: a systematic review. Journal of Neurodevelopmental Disorders, 11, 20. https://doi.org/10.1186/s11689-019-9279-8

29. Hansen, J.A. (2019). Development and psychometric evaluation of the Hansen Research Services matrix adaptive test: a measure of nonverbal IQ. J Autism Dev Disord, 49, 2721–2732. https://doi.org/10.1007/s10803-016-2932-0

30. Horwood, J., Salvi, G., Thomas, K., Duffy, L., Gunnell, D., Hollis, C., … Harrison, G. (2008). IQ and non-clinical psychotic symptoms in 12-year-olds: Results from the ALSPAC birth cohort. British Journal of Psychiatry, 193(*3*), 185–191. doi: https://10.1192/bjp.bp.108.051904

31. Iossifov, I., O’Roak, B. J., Sanders, S. J., Ronemus, M., Krumm, N., Levy, D., et al. (2014). The contribution of de novo coding mutations to autism spectrum disorder. Nature, 515, 216–221.

32. Krasileva, K. E., Sanders, S. J. & Bal, V. H. (2017). Peabody Picture Vocabulary Test: Proxy for Verbal IQ in Genetic Studies of Autism Spectrum Disorder. J Autism Dev Disord, 47, 1073–1085. https://doi.org/10.1007/s10803-017-3030-7

33. Kuhn, M. (2008). Building predictive models in R using the caret package. J Stat Softw, 28(*5*), 1–26.

34. Lombardo, M. V., Lai, M. C., Baron-Cohen, S. (2019). Big data approaches to decomposing heterogeneity across the autism spectrum. Mol Psychiatry, 24, 1435– 1450. https://doi.org/10.1038/s41380-018-0321-0

35. Mason, D., Capp, S. J., Stewart, G. R. et al. (2020). A meta-analysis of outcome studies of autistic adults: quantifying effect size, quality, and meta-regression. J Autism Dev Disord. https://doi.org/10.1007/s10803-020-04763-2

36. Maglogiannis, I. G. (Ed.). (2007). Emerging artificial intelligence applications in computer engineering: real word ai systems with applications in ehealth, hci, information retrieval and pervasive technologies (Vol. 160). Ios Press.

37. Mullen, E. M. (1995). Mullen scales of early learning (AGS ed.). Circle Pines: Pearson Assessment.

38. Munson, J., Dawson, G., Sterling, L., Beauchaine, T., Zhou, A., Elizabeth, K., Lord, C., Rogers, S., Sigman, M., Estes, A., & Abbott, R. (2008). Evidence for latent classes of IQ in young children with autism spectrum disorder. American journal of mental retardation: AJMR, 113(*6*), 439–452. https://doi.org/10.1352/2008.113:439-452

39. Nouwens, P. J. G., Lucas, R., Smulders, N. B. M., Ebregts, P. J. C. M., van Nieuwenhuizen, C. (2017). Identifying classes of persons with mild intellectual disability or borderline intellectual functioning: a latent class analysis. BMC Psychiatry, 17(*1*), 257.

40. Ogutu, J. O., Piepho, H. P., & Schulz-Streeck, T. (2011, December). A comparison of random forests, boosting and support vector machines for genomic selection. In BMC proceedings (Vol. 5, No. 3, pp. 1–5). BioMed Central.

41. Peyre, H., Charkaluk, M. L., Forhan, A., Heude, B., Franck, R. (2017). Do developmental milestones at 4, 8, 12 and 24 months predict IQ at 5–6 years old? Results of the EDEN mother–child cohort. European Journal of Paediatric Neurology, 21(2), 272-279.

42. Porter, J., Daniels, H., Feiler, A., Georgeson, J. (2011). Collecting disability data from parents, Research Papers in Education, 26(*4*), 427–443. DOI: 10.1080/02671520903281625

43. Rietvelt, C. A., Esko, T., Davies, G., et al. (2014). Common genetic variants and cognitive performance. Proceedings of the National Academy of Sciences, 111(*38*), 13790–13794. DOI: 10.1073/pnas.1404623111

44. Rauch et al. (2012). Range of genetic mutations associated with severe non-syndromic sporadic intellectual disability: an exome sequencing study. Lancet 380(*9854*),1674– 1682.

45. Reid, S.M., Meehan, E.M., Arnup, S.J. and Reddihough, D.S. (2018), Intellectual disability in cerebral palsy: a population-based retrospective study. Dev Med Child Neurol, 60: 687–694. https://doi.org/10.1111/dmcn.13773

46. Robinson, E. B, Samocha, K. E., Kosmicki, J., et al (2017). Autism spectrum disorder severity reflects the average contribution of de novo and familial influences. PNAS, 111 (*42*), 15161–15165.

47. Roid, G. (2003). Stanford Binet intelligence scales. Rolling Meadows, IL: Riverside Publishing.

48. Ronemus, M., Iossifov, I., Levy, D., Wigler, M. (2014). The role of de novo mutations in the genetics of autism spectrum disorders. Nat Rev Genet, 15(*2*), 133–141

49. Rutter, M., Bailey, A., Lord, C. (2003). The social communication questionnaire: manual. Western Psychological Services.

50. Samocha, K. E., Robinson, E. B., Sanders, S. J., et al. (2014). A framework for the interpretation of de novo mutation in human disease. Nat Genet, 46(*9*), 944–50. 10.1038/ng.3050.

51. Sanders, S. J., He, X., Willsey, A. J., Ercan-Sencicek, A. G., Samocha, K. E., Cicek, A. E., Murtha, M. T., Bal, V. H., Bishop, S. L., Dong, S., Goldberg, A. P., Jinlu, C., Keaney, J. F3rd., Klei, L., Mandell, J. D., Moreno-De-Luca, D., Poultney, C. S., Robinson, E. B., Smith, L., Solli-Nowlan, T., … State, M. W. (2015). Insights into Autism Spectrum Disorder Genomic Architecture and Biology from 71 Risk Loci. Neuron, 87(6), 1215– 1233. https://doi.org/10.1016/j.neuron.2015.09.016

52. Satterstrom, F. K., Kosmicki, J. A., Wang, J., Breen, M. S., De Rubeis, S., An, J. Y., Peng, M., Collins, R., Grove, J., Klei, L., Stevens, C., Reichert, J., Mulhern, M. S., Artomov, M., Gerges, S., Sheppard, B., Xu, X., Bhaduri, A., Norman, U., Brand, H., … Buxbaum, J. D. (2020). Large-Scale Exome Sequencing Study Implicates Both Developmental and Functional Changes in the Neurobiology of Autism. Cell, 180(*3*), 568–584.e23. https://doi.org/10.1016/j.cell.2019.12.036

53. Schoenberg, M. R., Lange, R. T., & Saklofske, D. H. (2007). A proposed method to estimate full scale intelligence quotient (FSIQ) for the Canadian Wechsler Intelligence Scale for Children–Fourth Edition (WISC–IV) using demographic and combined estimation procedures. Journal of Clinical and Experimental Neuropsychology, 29, 867– 878.

54. Scott, J..C, Van Pelt, A.E., Port, A.M., Njokweni, L., Gur, R.C., Moore, T.M., Phoi O., Tshume, O., Matshaba, M., Ruparel, K., Chapman, J., Lowenthal, E.D. (2020). Development of a computerised neurocognitive battery for children and adolescents with HIV in Botswana: study design and protocol for the Ntemoga study. BMJ Open, 10(8):e041099. doi: 10.1136/bmjopen-2020-041099.

55. Simonoff, E., Kent, R., Stringer, D., Lord, C., Briskman, J., Likito, S., Pickles, A., Charman, T., Baird, G. (2020). Trajectories in symptoms of autism and cognitive ability in autism from childhood to adult life: findings from a longitudinal epidemiological cohort. J Am Acad Child Adolesc Psychiatry, 59(*12*), 1342–1352.

56. SPARK Consortium (2018). SPARK: A US cohort of 50,000 families to accelerate autism research. Neuron, 97(*3*), 488–493. https://doi.org/10.1016/j.neuron.2018.01.015

57. Sparrow S.S., Cicchetti D.V. & Saulnier C.A. (2016). Vineland Adaptive Behavior Scales, Third Edition, San Antonio, TX, Pearson.

58. Stekhoven, D. J., & Bühlmann, P. (2012). MissForest—non-parametric missing value imputation for mixed-type data. Bioinformatics, 28(1), 112–118.

59. Stevens, E., Dixon, D. R., Novack, M. N., et al. (2019). Identification and analysis of behavioral phenotypes in autism spectrum disorder via unsupervised machine learning. Int J Med Inform, 129, 29–36.

60. Van Buuren, S., & Groothuis-Oudshoorn, K. (2011). mice: Multivariate imputation by chained equations in R. Journal of statistical software, 45(1), 1–67.

61. Veatch, O. J., Veenstra-VanderWeele, J., Potter, M., Pericak-Vance, M. A., Haines, J. L. (2014). Genetically meaningful phenotypic subgroups in autism spectrum disorders. *Genes*, Brain and Behavior, 13, 276–285 DOI: 10.1111/gbb.12117

62. Wang, T., Guo, H., Xiong, B., Stessman, H. A., Wu, H., Coe, B. P., Turner, T. N., Liu, Y., Zhao, W., Hoekzema, K., Vives, L., Xia, L., Tang, M., Ou, J., Chen, B., Shen, Y., Xun, G., Long, M., Lin, J., Kronenberg, Z. N., … Eichler, E. E. (2016). De novo genic mutations among a Chinese autism spectrum disorder cohort. Nature communications, 7, 13316. https://doi.org/10.1038/ncomms13316

63. Warnell, F., George, B., McConachie, H., Johnson, M., Hardy, R., & Parr, J. R. (2015). Designing and recruiting to UK autism spectrum disorder research databases: do they include representative children with valid ASD diagnoses?. BMJ open, 5(*9*), e008625. https://doi.org/10.1136/bmjopen-2015-008625

64. Wechsler, D. (2014). Wechsler intelligence scale for children (5^th^ ed.). Bloomington, MN: NCS Pearson.

65. Wilson, B.N., Kaplan, B.J., Crawford, S.G., Campbell, A., Dewey, D. (2000). Reliability and validity of a parent questionnaire on childhood motor skills. Am J Occup Ther 54(*5*): 484–493.

